# Convolutional Neural Networks for Burn Segmentation and Classification Tasks Using RGB Photographs: A Five-Year Systematic Review

**DOI:** 10.1101/2025.07.08.25331144

**Authors:** Yu-Tung Fu

## Abstract

**Background:** Burn wound assessment remains complex, with visual accuracy often below 50% among non-specialists. Convolutional neural networks (CNNs) offer promising solutions, achieving 68.9%–95.4% accuracy in depth classification and 76.0%–99.4% in area segmentation. This review systematically evaluates CNN-based burn area segmentation (BAS), burn depth classification (BDC), and burn depth segmentation (BDS) using RGB photographs.

**Methods:** A systematic search of PubMed, Medline, Embase, and Cochrane Library (January 2020–April 2025) was conducted on 1st April 2025. Studies applying CNNs to RGB images for one or more of the prediction tasks (BAS, BDC, or BDS) were included. Risk of bias was assessed using PROBAST+AI. Data on model architecture, datasets, and performance metrics were extracted and synthesized narratively.

**Result:** A total of 14 studies were included. Among BAS tasks, six reported accuracy above 90%, and five reported Dice Coefficient over 0.8. The combination of ResNet-101 and ASPP provides a strong and stable baseline across studies. In BDC tasks, four reported an accuracy above 80%, and all six reported an F1 score above 0.73. The top two best-performing models employed feature enhancement strategies to achieve accuracy up to 98% and F1 score of 0.97. In BDS tasks, low-quality data and inconsistent annotation were observed to negatively affect model performance.

**Conclusion:** CNN-based models show strong potential in burn wound analysis using RGB images. However, considerable heterogeneity remains, and future studies should prioritise head-to-head comparisons and multicentre validation to strengthen model generalisability.

## 1. Introduction

Burn injuries remain a significant public health concern worldwide, affecting both adults and children across diverse healthcare settings. In the United States alone, it is estimated that approximately 500,000 individuals seek medical care for burn-related injuries annually(1). In addition to the physical harm, burn injuries frequently lead to considerable morbidity, compromised psychological well-being, reduced quality of life, and substantial financial burden. For example, in Spain, the average annual healthcare cost per burn patient has been estimated at approximately USD 99,773(2, 3). In many low-resource settings, however, the impact of burn injuries may be underestimated due to limited access to medical care and underreporting.

Among the entire process of burn care, precise evaluation of burn severity and total body surface area (TBSA) is critical, as it plays a vital role in guiding resuscitation, surgical planning, and ultimately, patient survival(4, 5). At present, visual assessment remains the most commonly used and accessible method for evaluating burn injuries. However, its accuracy has been reported to range between 66% and 75% among highly experienced burn specialists, and falls below 50% in less experienced practitioners, suggesting that this method may not always provide consistent or reliable results (6, 7). Laser Doppler Imaging (LDI) represents an alternative modality, renowned for its high accuracy in assessing burn depth. When infected wounds are excluded, LDI has demonstrated an accuracy of up to 99% (8) and a sensitivity of 91% in predicting wound healing time (9). Despite these advantages, its widespread application is hindered by high cost, lack of portability, and relatively low efficiency, rendering it impractical for use in smaller or remote healthcare facilities(10, 11).

Given the aforementioned limitations, deep learning techniques, especially Convolutional Neural Networks (CNNs), have shown substantial promise and are considered one of the most potent technologies for automated wound segmentation and burn severity classification(12). CNNs have become widely adopted in recent years due to their rapid and effective feature extraction, particularly for subtle features such as edges, margins, and textures that may be missed by other machine learning architectures. This capability is particularly advantageous for burn-related tasks that require fine-grained spatial recognition of wound boundaries and depth. According to Taib et al.(13), a systematic review of studies employing artificial intelligence (AI) for burn assessment reported that the accuracy of AI-based burn depth classification ranged from 68.9% to 95.4%, while that of burn segmentation ranged from 76.0% to 99.4%. The precision of such predictions facilitates rapid communication among specialists, thereby supporting timely decisions regarding fluid resuscitation requirements or the need for surgical intervention.

While Taib et al.’s study included heterogeneous study designs, machine learning models, and data types spanning from 1990 to 2023, this present review focuses specifically on CNN-based models applied to RGB images, a more practical and accessible imaging format in both hospital and low-resource environments. In addition, this review adopts a task-based classification of CNN applications in burn image analysis, distinguishing three prediction tasks: burn area segmentation (BAS), burn depth classification (BDC), and burn depth segmentation (BDS). To our knowledge, no previous review has systematically examined CNN-based models specifically for RGB burn images, nor introduced a task-based categorisation such as BAS, BDC, and BDS. By synthesising current evidence, this review aims to evaluate the performance and limitations of CNN-based models and to clarify the clinical applicability of RGB images in burn wound assessment.

## 2. Method

This systematic review was conducted following the PRISMA guidelines (Preferred Reporting Items for Systematic Reviews and Meta-Analyses).

### 2.1 Search Strategy

To ensure both comprehensive coverage and precision, a balanced search strategy was designed to optimise sensitivity and specificity. A systematic search was conducted on 1^st^ April 2025 by the author alone across multiple databases, including MEDLINE, EMBASE (Excerpta Medica Database), PubMed, and the Cochrane Library. Although the Cochrane Library was included for completeness, no eligible studies were retrieved from this database. The core Boolean string used in PubMed was: ((“burn segmentation”) OR (“burn depth”)) AND ((“artificial intelligence”) OR (“machine learning”) OR (“convolutional neural networks” OR “CNN”))

In addition, Medical Subject Headings (MeSH) such as “Burns"[MeSH] and “Artificial Intelligence"[MeSH] were incorporated where applicable. Equivalent adaptations were applied to EMBASE, MEDLINE, and the Cochrane Library using their respective thesaurus systems.

The search was limited to studies published in English between 1 January 2020 and 1 April 2025. Additionally, one article was identified through manual reference screening.

### 2.2 Eligibility Criteria

Studies were included if they: (1) used standard RGB photographic images as input data; (2) applied convolutional neural networks (CNNs); (3) addressed one or more of the following tasks: burn area segmentation, burn depth classification, or burn depth segmentation; (4) were published between 1 January 2020 and 1 April 2025; and (5) were available in full-text English.

Studies were excluded if they: (1) did not use CNNs; (2) used non-photographic imaging modalities such as multispectral imaging or Laser Doppler Imaging (LDI); or (3) were not published in English or lacked full-text availability.

Studies that employed multimodal data (e.g., RGB and ultrasound) were included if the performance metrics for RGB-only input were reported and could be separately analysed.

### 2.3 Definition of prediction tasks

The current studies utilizing CNNs for burn wound assessment have adopted heterogeneous prediction tasks, making it essential to clearly define these prediction tasks for a comprehensive analysis and better understanding. This review identified three primary prediction tasks: (1) burn area segmentation, (2) burn depth classification, and (3) burn depth segmentation.

Burn area segmentation (BAS) refers to the identification and delineation of the burn margin, regardless of the depth of the wound. CNN models performing this task are trained to classify each pixel as either “burn” or “non-burn,” further delineating the entire margin of the burn wound. Studies utilising object detection as the prediction task were excluded from this category, as their models only label coarse regions rather than performing pixel-level margin delineation.

Burn depth classification (BDC) primarily aims to distinguish subtle differences among various depth levels (e.g. superficial-dermal, deep-dermal, and full-thickness). This task typically produces a single depth label for an entire image or a specific burn region. However, as it does not provide pixel-wise depth information, this approach may be insufficient for applications requiring detailed spatial assessment of burn severity.

Burn depth segmentation (BDS), in contrast, predicts depth categories at the pixel level. In this task, CNN models classify each pixel as either a specific burn depth (e.g. superficial-dermal, deep-dermal, or full-thickness) or background (non-burn area). Upon completing the full prediction process, the models enable delineation of the wound margin and visualisation of depth distribution, offering a more comprehensive representation of burn severity.

Some studies included more than one prediction task, and the performance of each task was analysed separately, as summarised in Table 1.

**Table 1.**
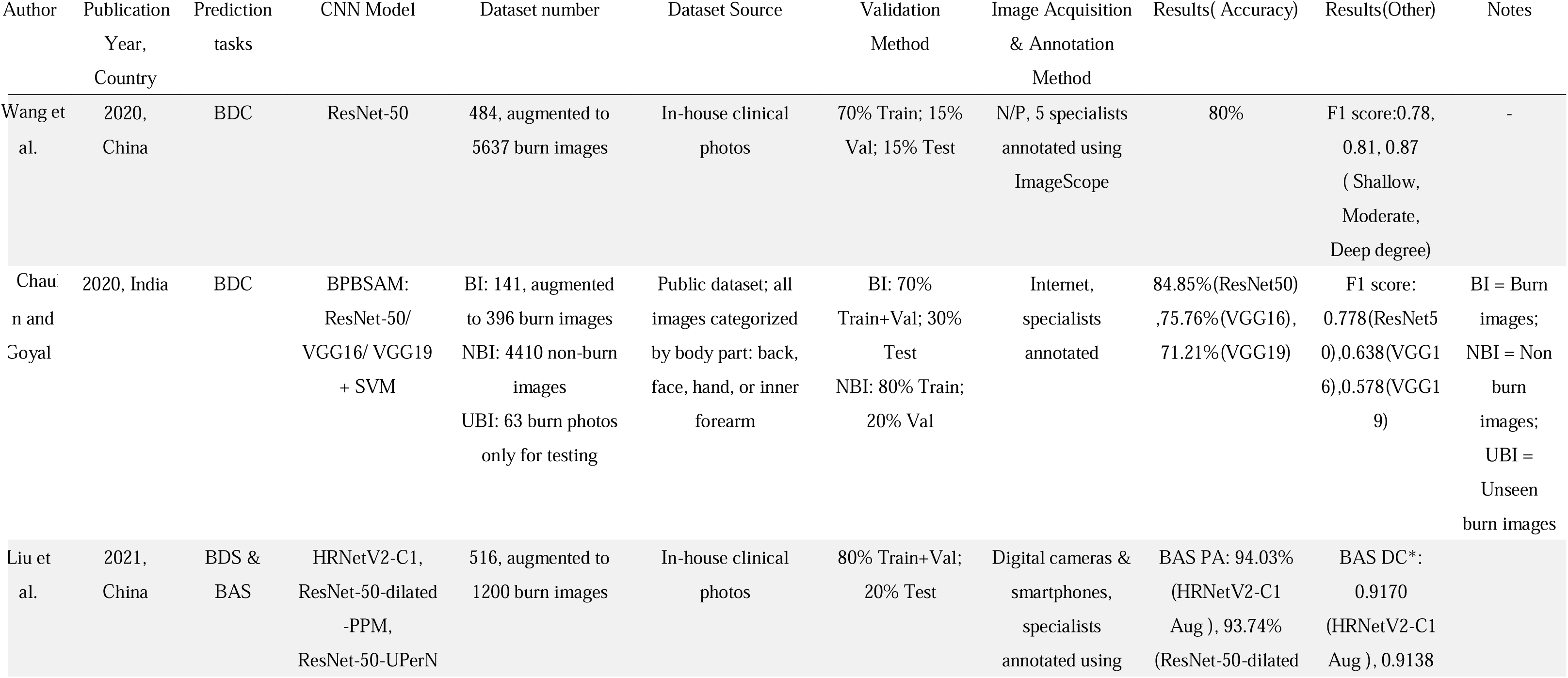
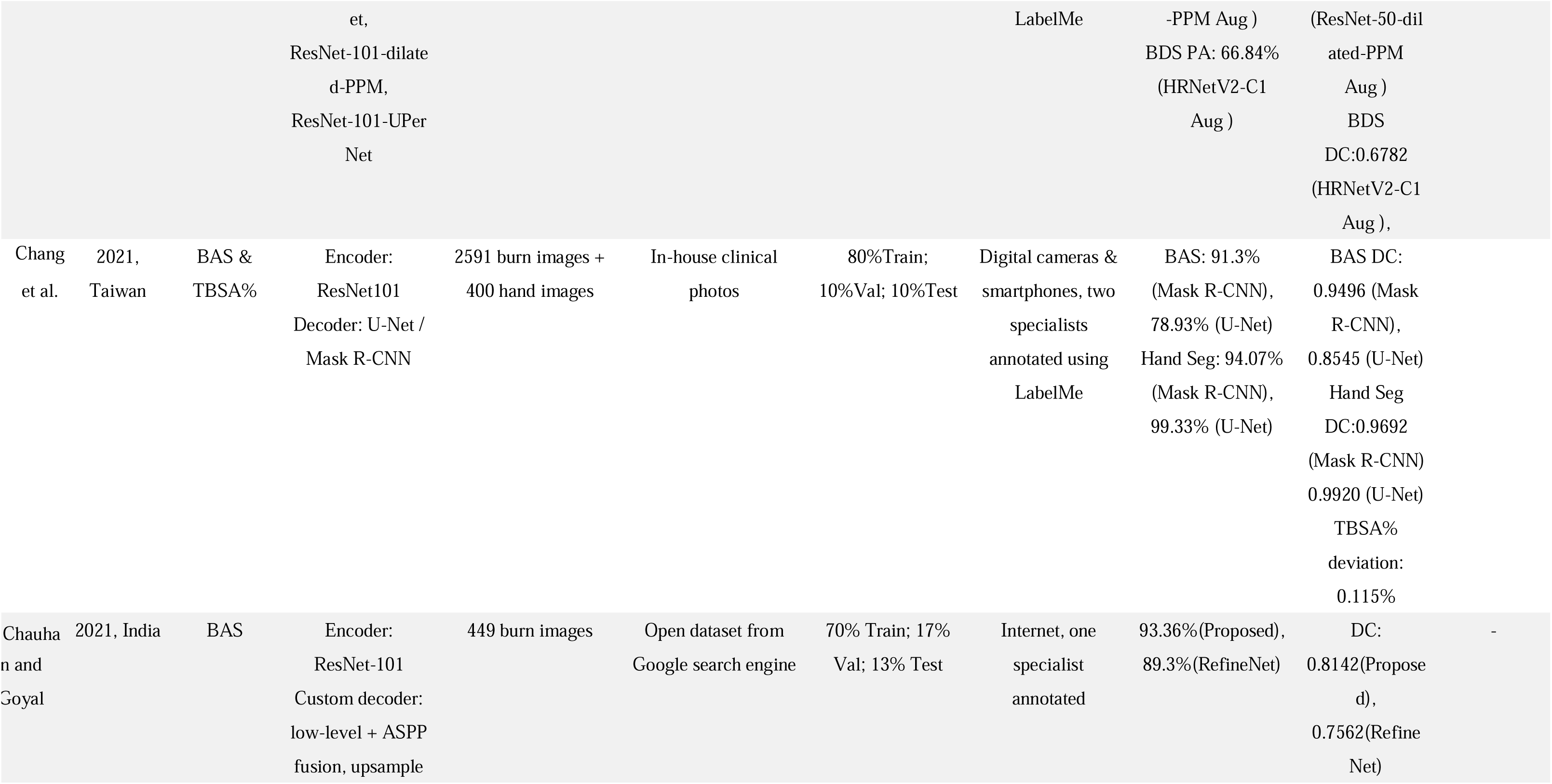
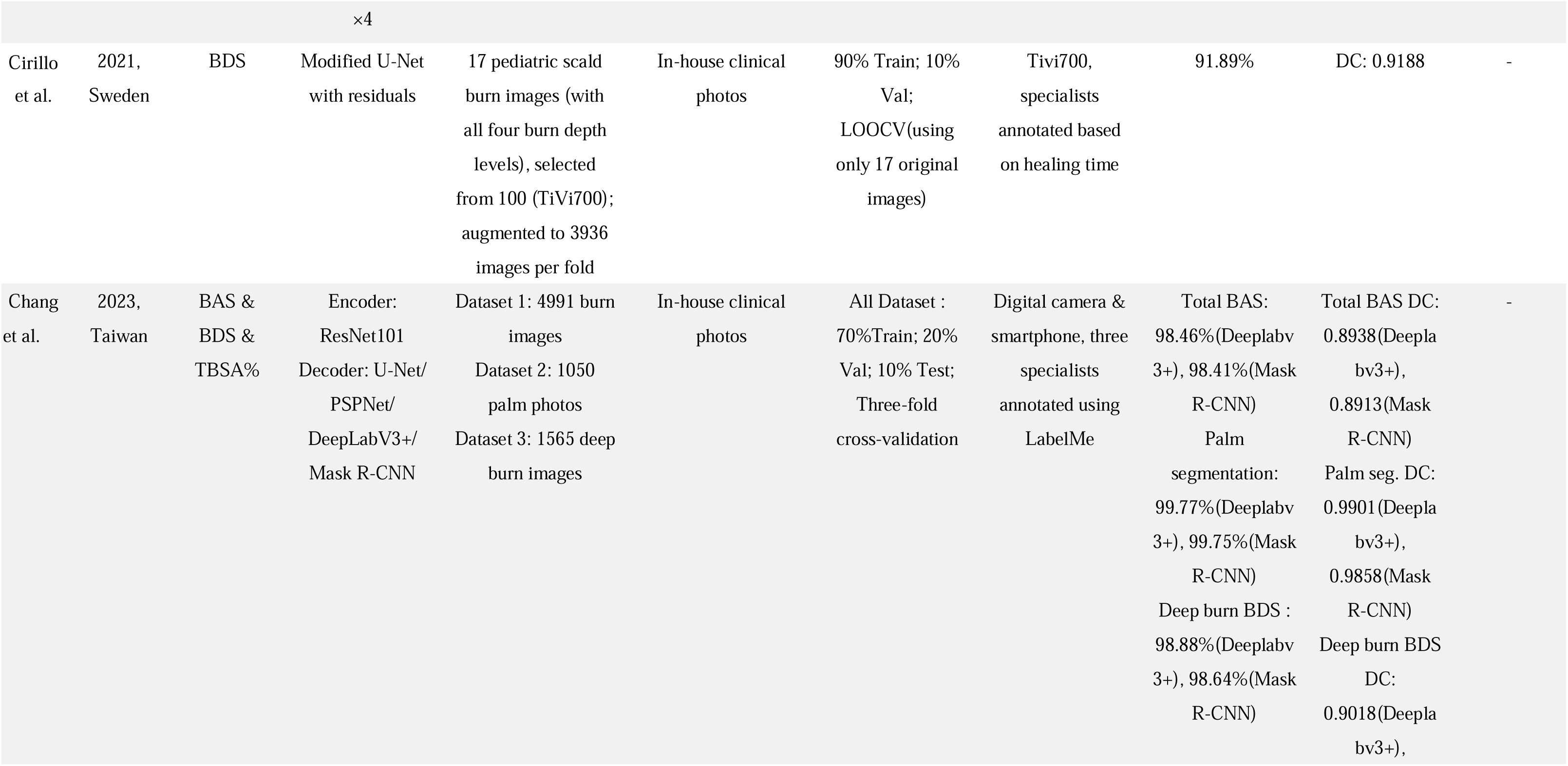
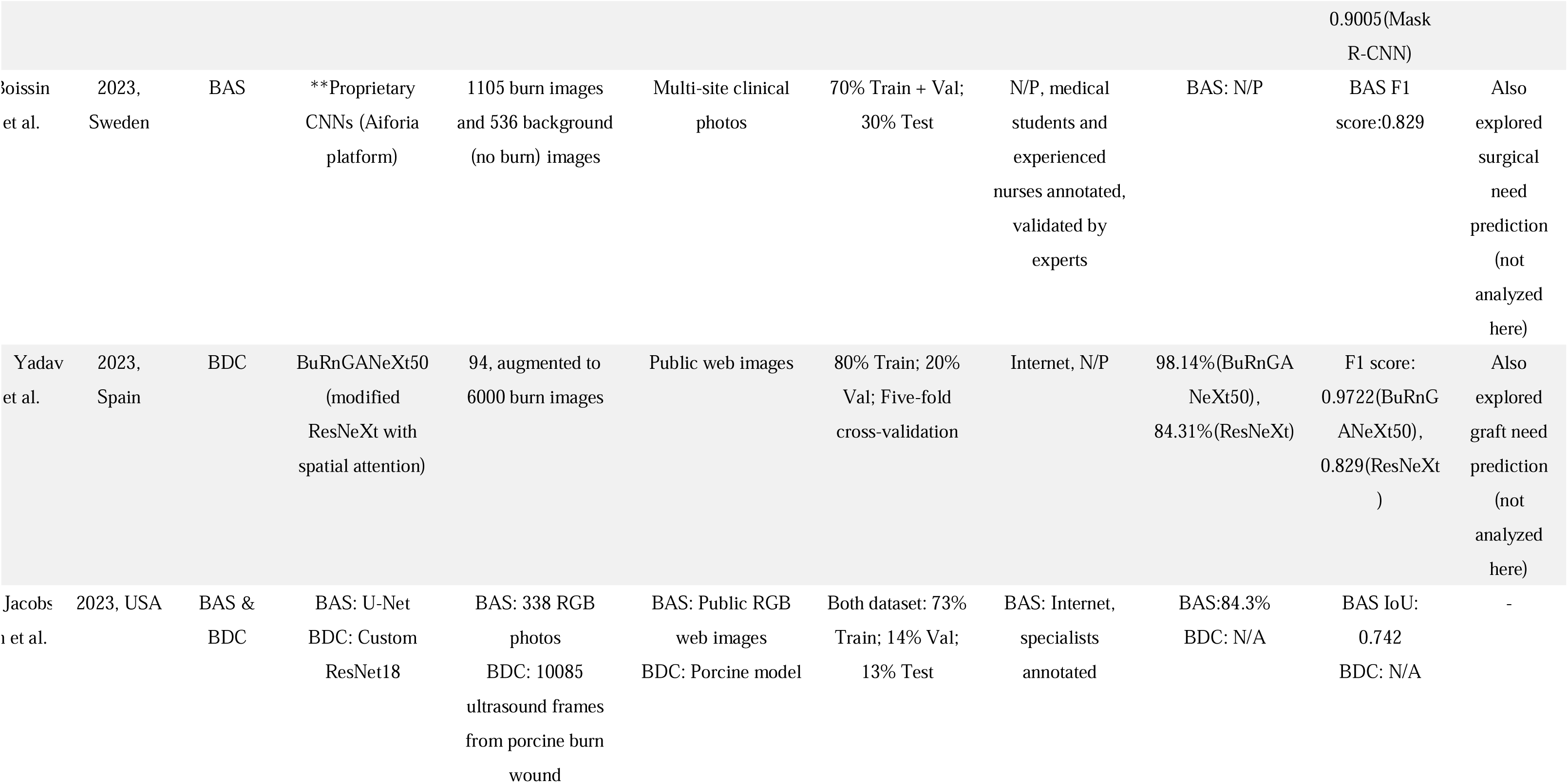
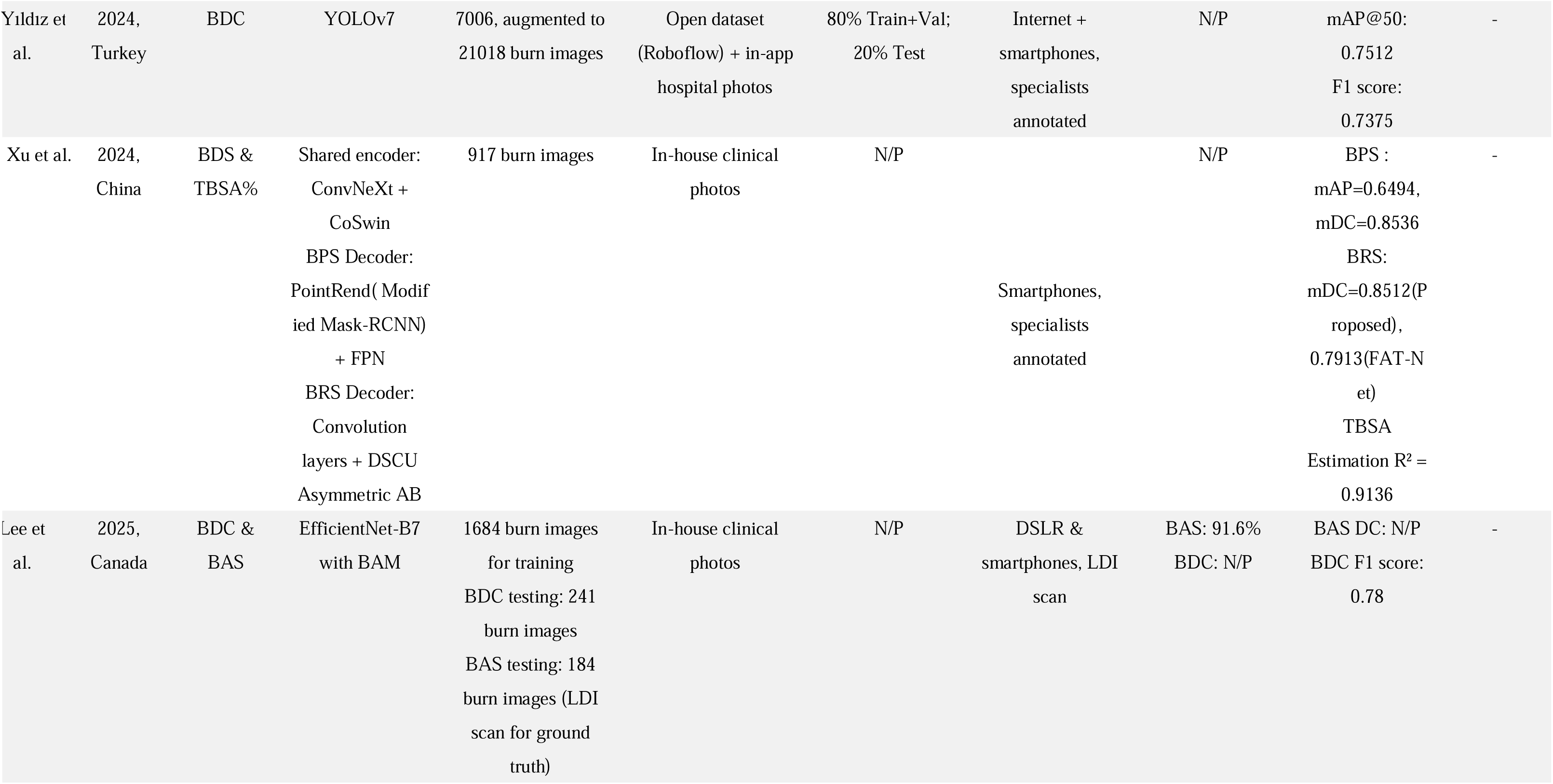
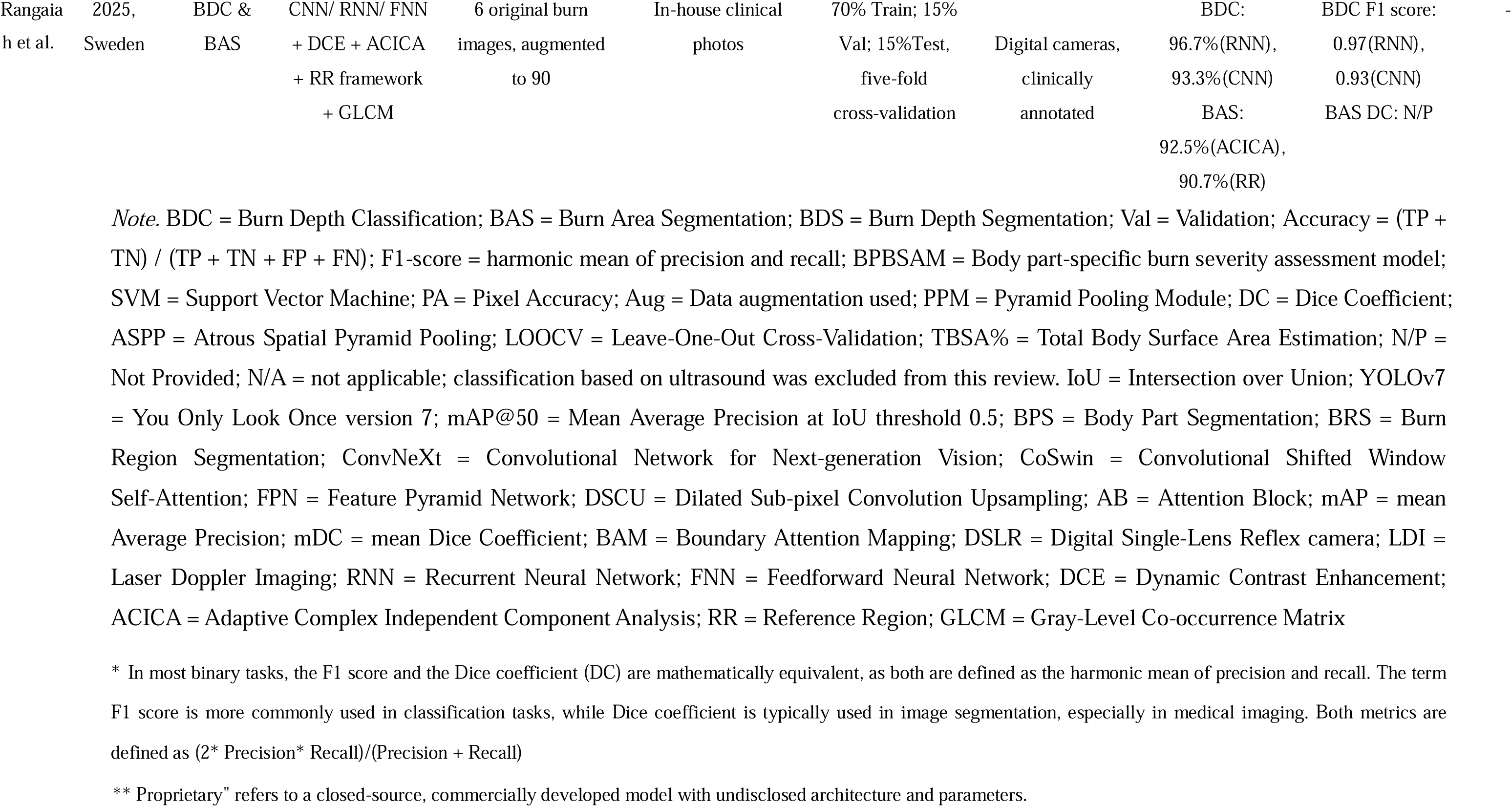
Summary of included studies on CNN-based models for burn wound assessment, including prediction tasks, datasets, model architectures, validation methods, and performance metrics.

### 2.4 Evaluation Metrics

Given the nature of the prediction tasks, different performance metrics were commonly used across studies. For segmentation tasks, including BAS and BDS, accuracy and the Dice coefficient (DC) were most frequently reported. The former indicates the proportion of correctly classified pixels across the entire image, whereas the latter measures the overlap between predicted and ground truth masks at the pixel level.

In contrast, BDC, a classification task typically producing a single label per image or region, was more commonly evaluated using F1 score, which balances precision and recall and is suitable for imbalanced data. Accuracy was also reported for this task reflecting the proportion of correctly classified burn depth categories among all samples. Additional data items extracted included sample size, source of dataset (e.g., clinical or non-clinical), model architectures used. No assumptions were made for missing or unclear information.

### 2.5 Data synthesis

Given the expected methodological and outcome heterogeneity among included studies, a narrative synthesis was conducted. Studies were grouped according to their prediction tasks (BAS, BDC, BDS). Due to substantial variability in study designs and performance metrics, meta-analysis was not performed.

### 2.6 Risk of Bias Assessment

Data extraction and risk of bias assessment were conducted independently by the author.

The risk of bias (RoB) for each included study was assessed using the PROBAST+AI tool(14), a tailored extension of the original PROBAST instrument for prediction model studies incorporating artificial intelligence. Four key domains were evaluated: Participants, Predictors, Outcomes, and Analysis. Based on domain-level assessments, each study was categorised as having low, moderate, or high overall risk of bias. Studies using non-clinical datasets (e.g., Google Images or iStock photos) or lacking consistent annotation protocols were considered at high risk in the Participants or Outcome domains. Full RoB assessments are provided in the supplementary table 1.

### 2.7 Other Information

This systematic review was not registered in any publicly accessible database. No formal protocol was prepared prior to conducting the review. Consequently, no amendments to a registered record or protocol were applicable. The PRISMA 2020 Checklist is provided in Supplementary Table 2.

## 3. Results

A total of 115 articles were initially identified through the database search. After removing duplicates, 97 records remained for title and abstract screening. Of these, 79 were excluded based on relevance, resulting in 18 studies selected for full-text review. Two of these were subsequently excluded due to the unavailability of full-text access. Among the 16 full-text articles assessed, two were excluded: one due to a mismatch in research focus(15), and the other due to an outcome not related to segmentation or classification(16). Details of excluded full-text studies are provided in Supplementary Table 3. A PRISMA flow diagram was constructed to illustrate the study selection process (Figure 1). In total, 14 studies met the inclusion criteria and were included in this review.(17-30)

**Figure 1.**
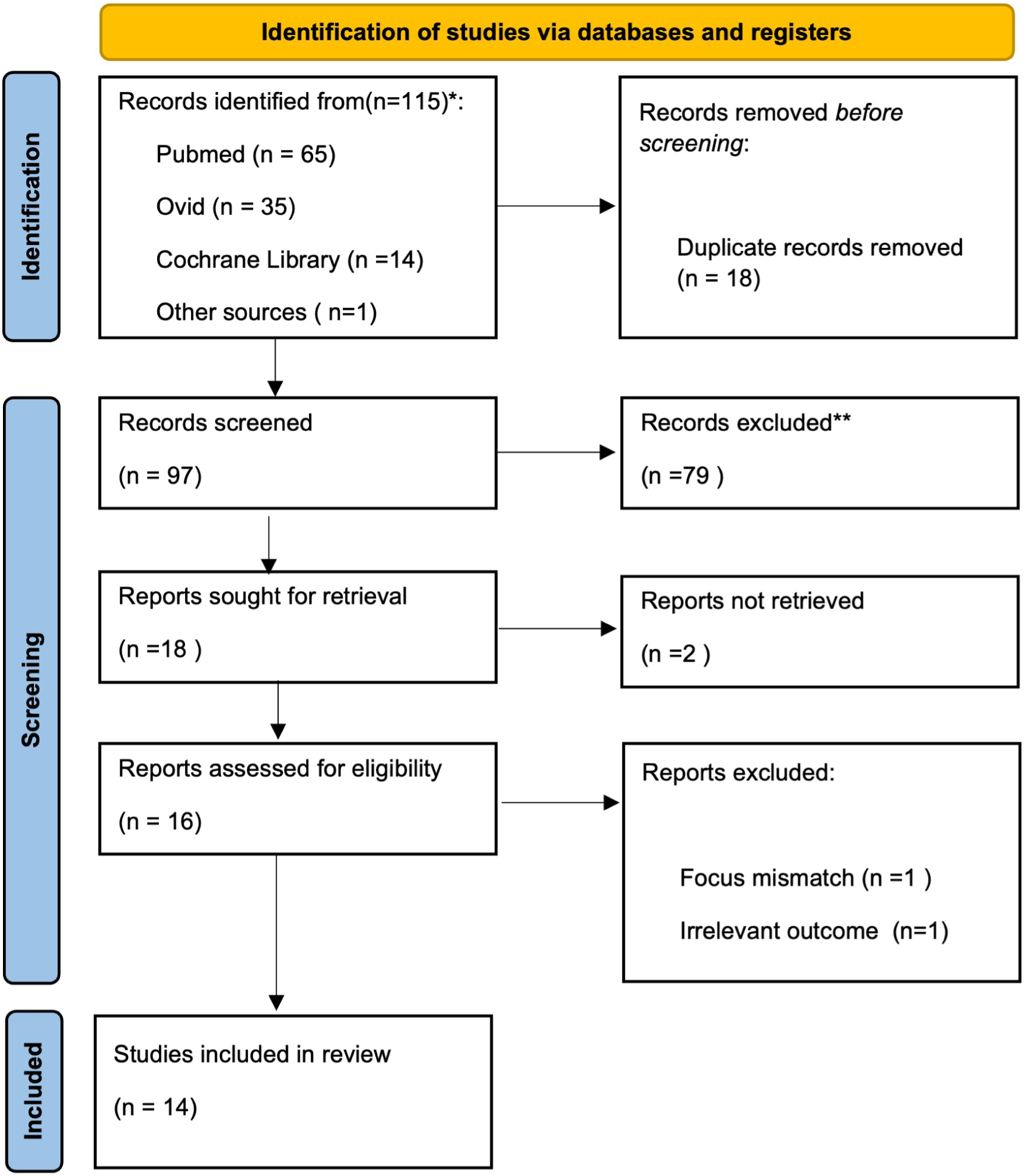
PRISMA diagram. The figure illustrates the study selection process, including identification, screening, eligibility, and final inclusion.

### 3.1 Risk of bias results

Among the fourteen studies, four were assessed as having a low risk of bias, four as moderate risk, and six as high risk. Notably, the most frequent sources of bias were observed in the Participants domain, where five of the six studies assessed as high risk were classified as such due to the use of public datasets for training or validation, which did not meet the criteria for low risk in PROBAST+AI. These datasets failed to adequately represent the target population in real-world settings and lacked systematic and consistent data collection procedures, thereby introducing a high risk of selection bias. Additionally, most studies classified as having moderate risk in the Outcome domain were downgraded due to a lack of transparency in expert annotation or ground truth labelling, thereby increasing uncertainty regarding the gold standard used for training. Figure 2 shows the distribution of overall risk of bias, and Figure 3 presents the domain-level risk assessments (D1–D4) across the included studies.

**Figure 2.**
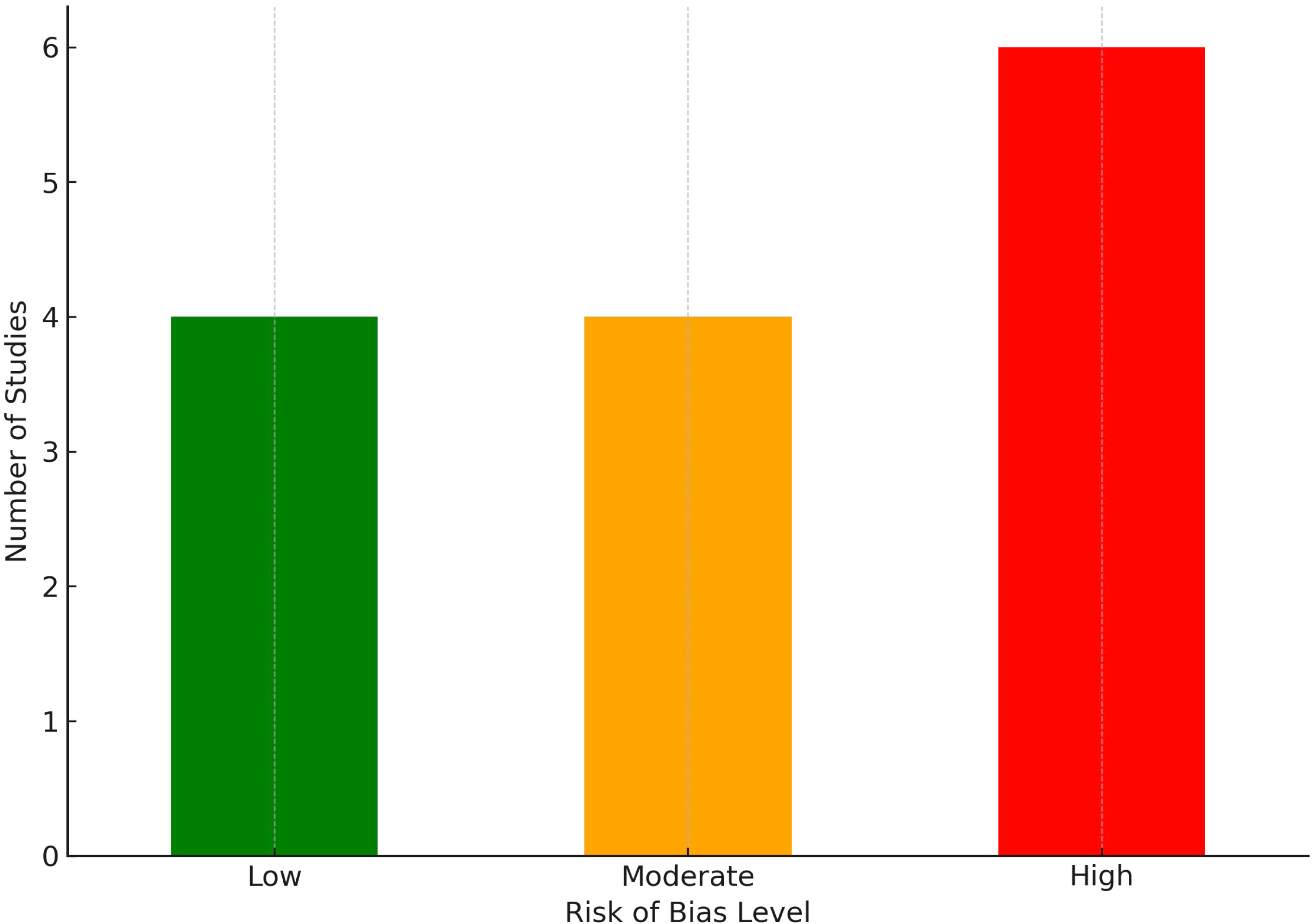
Overall risk of bias distribution across included studies.

**Figure 3.**
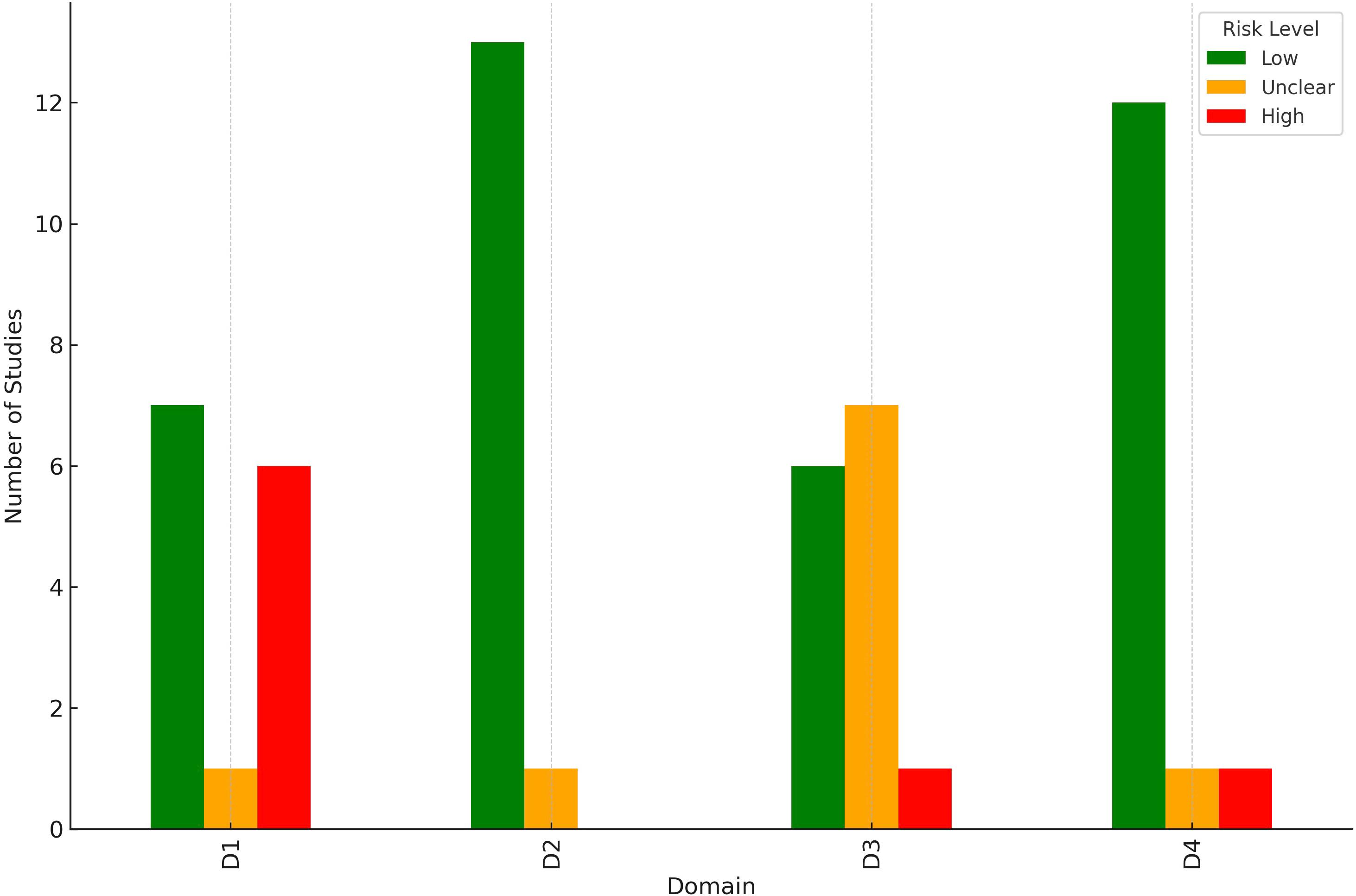
Risk of bias by domain (D1 = Participants, D2 = Predictors, D3 = Outcomes, D4 = Analysis).

### 3.2 Burn Area Segmentation (BAS)

Eight out of the fourteen studies included in this review performed a burn area segmentation task as part of their methodology(19-21, 23, 24, 26, 29, 30). The corresponding model performance is summarised in Table 1.

Across all eight studies, accuracy ranged from 84.3% to 98.46%, with six studies reporting values above 90%. Dice coefficient (DC) ranged from 0.81 to 0.95, with five studies reporting values above 0.8, suggesting a generally strong overlap between predicted and ground truth masks.

*DeepLabV3+ with ResNet-101 backbone*, developed by Chang et al.(2023), and *HRNetV2-C1 Aug*, developed by Liu et al., achieved the best performance in BAS, reporting an accuracy of 98.46% and a DC of 0.8938, and an accuracy of 94.03% and a DC of 0.9170, respectively.

The third-best models included *ResNet-101 with Atrous Spatial Pyramid Pooling (ASPP)*, developed by Chauhan and Goyal (2021) (accuracy: 93.36%, DC: 0.8142), and *Mask R-CNN with ResNet-101 backbone*, developed by Chang et al.(2021) (accuracy: 91.3%, DC: 0.95).

Notably, three(20, 21, 23) of the top four best-performing studies employed *ResNet-101* as the encoder, with two(21, 23) also incorporated *Atrous Spatial Pyramid Pooling (ASPP)*. In contrast, the three lowest-performing studies did not utilise either of these architectures components. This pattern highlights the potential utility of the combination of *ResNet-101 and ASPP* as a strong and reliable baseline for BAS in CNN-based models.

### 3.3 Burn Depth Classification (BDC)

Among the fourteen studies included in this review, six conducted burn depth classification as one of their outcome measures(17, 18, 25, 27, 29, 30). See Table 1 for model performance metrics. Among all six studies, accuracy ranged from 80% to 98.14%, and F1 score ranged from 0.74 to 0.97. *BuRnGANeXt50*, developed by Yadav et al., achieved the best performance, with an accuracy of 98.14% and an F1 score of 0.9722. The second-best model was a *Recurrent Neural Network (RNN) with multiple image preprocessing steps*, developed by Rangaiah et al., which achieved an accuracy of 96.7% and an F1 score of 0.97. *ResNet-50* was used in both the studies by Chauhan and Goyal (2020) (accuracy: 84.85%, F1 score: 0.78), and by Wang et al. (accuracy: 80%, F1 score: 0.82), and ranked third overall in terms of performance. Despite some variation in architecture and preprocessing strategies, most models achieved moderate to high performance in BDC.

Notably, the two highest-performing models(25, 30) both employed feature enhancement strategies, which may have contributed to the 13% accuracy and 0.19 F1 score gain over the third-ranked model, suggesting that feature enhancement may play a role in boosting classification performance.

### 3.4 Burn Depth Segmentation (BDS)

Only four out of fourteen studies employed burn depth segmentation as one of their prediction tasks(19, 22, 23, 28). The details of these models are listed in Table 1. *DeepLabV3+*, developed by Chang et al.(2023), achieved the highest accuracy (98.88%) and the second-highest DC of 0.9018, whereas the *Modified U-Net with residual connections*, developed by Cirillo et al., reported the highest DC of 0.9188 and the second-highest accuracy (91.89%). For the remaining two studies, while Xu et al.’s model still achieved a relatively high DC of 0.8512, Liu et al.’s model (*HRNetV2-C1 Aug*) reported a notably lower accuracy of 66.84% and a DC of 0.6782, highlighting the heterogeneity in performance across studies.

Notably, while both Cirillo et al. and Liu et al. employed advanced CNN-based semantic segmentation architectures, the 25% difference in accuracy and 0.24 disparity in DC may reflect factors beyond model design, such as dataset quality or annotation consistency, which will be detailed in Discussion.

## 4. Discussion

It is important to note that the included studies employed heterogeneous study designs, dataset characteristics, and annotation quality. These factors may have contributed to the significant differences among performance metrics across CNN models, making cross-study comparisons more challenging. Nevertheless, this review highlights several meaningful performance trends, suggesting which architectural components may be most effective for burn wound assessment.

### 4.1 Burn Area Segmentation (BAS)

To support the interpretation of burn area segmentation performance, Table 2 and Figure 4 summarise the key metrics of the included CNN models. Among the top four best-performing studies, three(20, 21, 23) employed *ResNet-101* as the encoder, with two(21, 23) also incorporating *Atrous Spatial Pyramid Pooling (ASPP)*. Notably, the *DeepLabV3+* model developed by Chang et al.(2023), corresponding to study 21, also included *ASPP* in its architecture. The strong performance of the combination of *ResNet-101 with ASPP* is not unexpected, as *ResNet-101*, used as the backbone, is well known for its strong feature extraction capabilities through deep residual learning and skip connections. These architectural features allow the model to retain information across layers and to learn high-level abstract semantic representations, thereby enhancing model performance on classification tasks(31, 32). Furthermore, *ASPP* is specifically designed for effective multiscale feature extraction, as it expands the receptive field while preserving spatial resolution. This enables *ASPP* to capture both fine-grained details and broader contextual information within an image, thereby enhancing performance in complex tasks such as semantic segmentation(33-35). The combination of *ResNet-101 and ASPP* is particularly well-suited for tasks requiring precise boundary delineation across objects of varying sizes, such as burn wounds.

**Figure 4.**
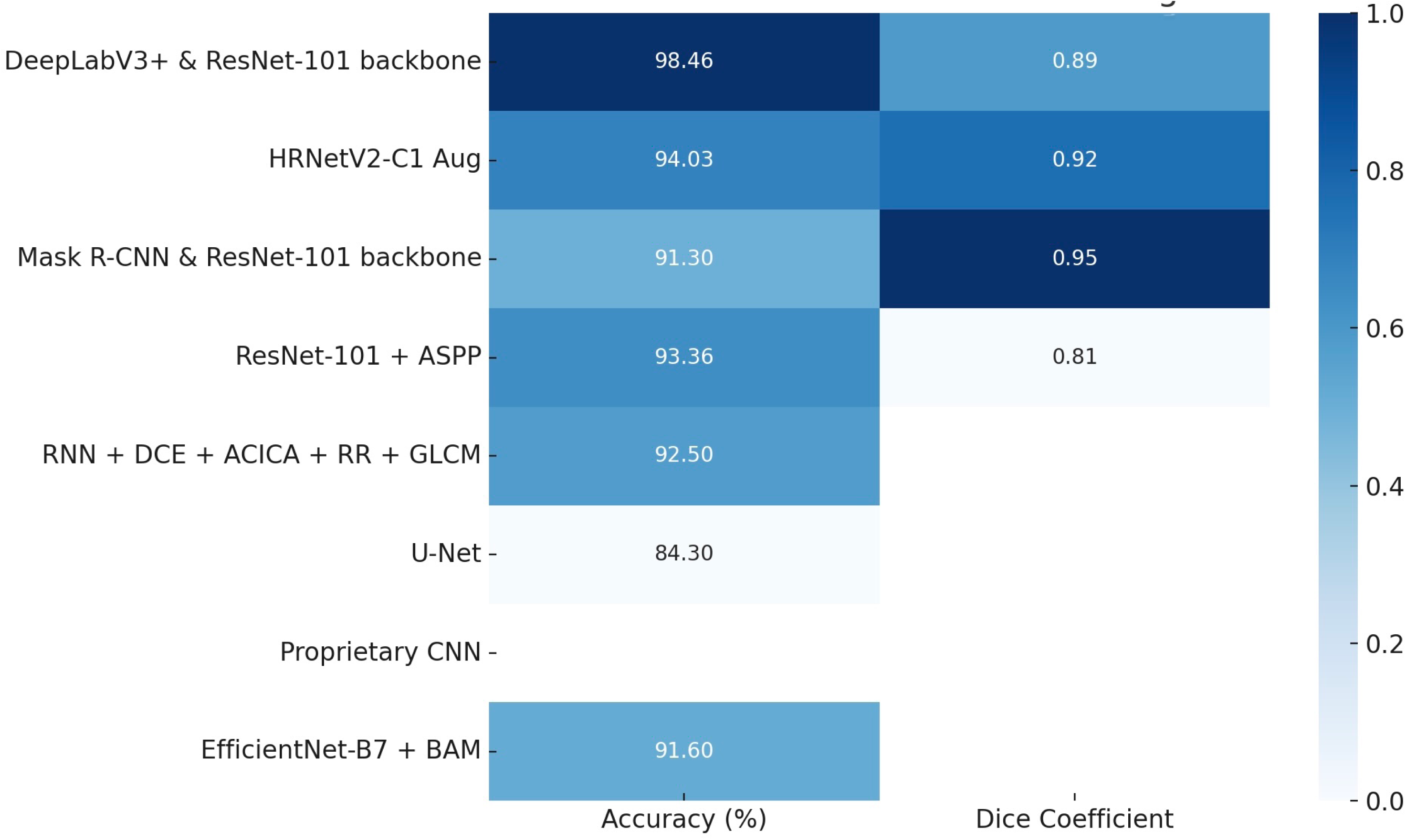
Normalised heatmap of CNN model performance on burn area segmentation (BAS). This figure presents the relative performance of selected CNN models in terms of accuracy and Dice coefficient. Darker shades indicate higher normalized values. Models with missing data are shown with gaps.

**Table 2.**
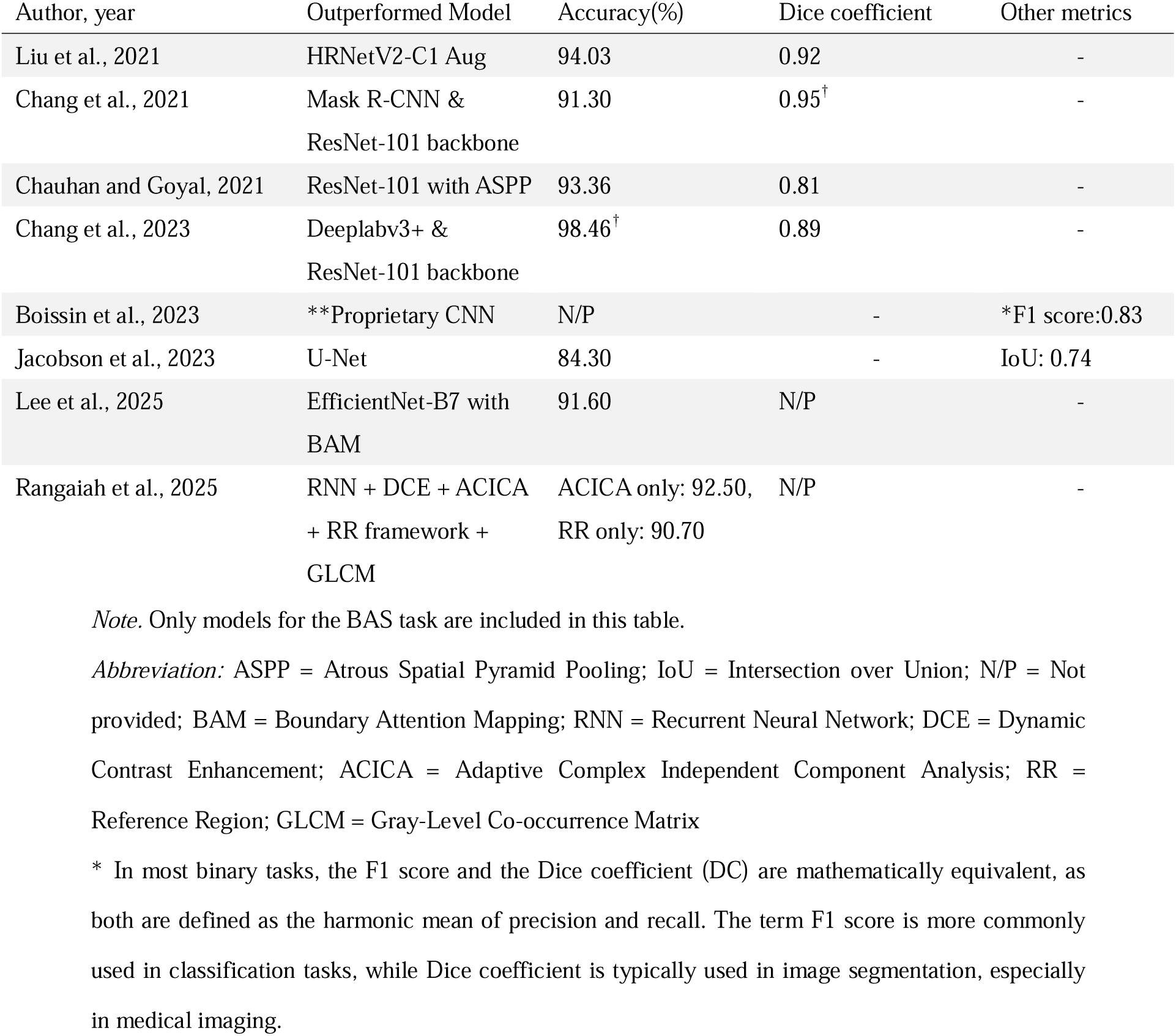
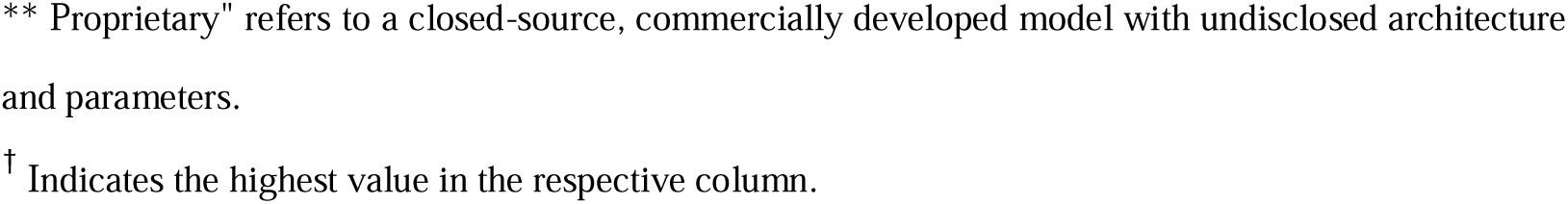
Summary of CNN Models for BAS: Accuracy and Complementary Performance Metrics.

| Author, year | Outperformed Model | Accuracy(%) | Dice coefficient | Other metrics |
| --- | --- | --- | --- | --- |
| Liu et al., 2021 | HRNetV2-C1 Aug | 94.03 | 0.92 | - |
| Chang et al., 2021 | Mask R-CNN & ResNet-101 backbone | 91.30 | 0.95<sup>†</sup> | - |
| Chauhan and Goyal, 2021 | ResNet-101 with ASPP | 93.36 | 0.81 | - |
| Chang et al., 2023 | Deeplabv3+ & ResNet-101 backbone | 98.46<sup>†</sup> | 0.89 | - |
| Boissin et al., 2023 | **Proprietary CNN | N/P | - | *F1 score:0.83 |
| Jacobson et al., 2023 | U-Net | 84.30 | - | IoU: 0.74 |
| Lee et al., 2025 | EfficientNet-B7 with BAM | 91.60 | N/P | - |
| Rangaiah et al., 2025 | RNN + DCE + ACICA + RR framework + GLCM | ACICA only: 92.50, RR only: 90.70 | N/P | - |

*Note.* Only models for the BAS task are included in this table.
*Abbreviation:* ASPP = Atrous Spatial Pyramid Pooling; IoU = Intersection over Union; N/P = Not provided; BAM = Boundary Attention Mapping; RNN = Recurrent Neural Network; DCE = Dynamic Contrast Enhancement; ACICA = Adaptive Complex Independent Component Analysis; RR = Reference Region; GLCM = Gray-Level Co-occurrence Matrix
\* In most binary tasks, the F1 score and the Dice coefficient (DC) are mathematically equivalent, as both are defined as the harmonic mean of precision and recall. The term F1 score is more commonly used in classification tasks, while Dice coefficient is typically used in image segmentation, especially in medical imaging.
\*\* Proprietary" refers to a closed-source, commercially developed model with undisclosed architecture and parameters.
<sup>†</sup> Indicates the highest value in the respective column.

However, it is important to note that in the study by Liu et al.(19), *HRNetV2-C1* outperformed *ResNet-101* as an encoder in terms of both accuracy and DC. Notably, this was the only study in the review that provided a direct head-to-head comparison of the two architectures within the same experimental setting. This may be due to *HRNetV2-C1*’s ability to maintain high-resolution feature representations throughout the processing stages and perform multi-scale fusion without downsampling. These architectural features are particularly effective in capturing complex structural variations, especially in low-resolution images, such as burn wounds or photographs captured by smartphones(36). In summary, the combination of *ResNet-101 and ASPP* provides a strong and stable baseline across studies, enabling models to achieve an accuracy above 93% and a DC of 0.81 on BAS task. However, *HRNetV2-C1* may be more suitable when precise spatial detail is required or when low-resolution data is employed. Further head-to-head comparative studies are needed to validate the relative performance of these two architectures.

### 4.2 Burn Depth Classification (BDC)

The models developed by Yadav et al. and Rangaiah et al., which achieved the best performance in this task, both adopted feature enhancement strategies to guide the models to focus on extracting important features or to enhance contrast and texture details in burn wound images, as shown in Table 3 and Figure 5. Yadav et al.’s model incorporated both spatial and channel attention modules. Channel attention helps the model prioritise the most relevant features by adjusting the significance of each channel, while spatial attention guides it to focus on important regions in the image, such as lesion borders and blisters(37). These mechanisms can significantly improve performance in burn depth classification, as they enable the model to distinguish between different depth degrees by extracting subtle yet important features. Rangaiah et al.’s study employed multiple image preprocessing techniques, such as DCE (Dynamic Contrast Enhancement), ACICA (Adaptive Complex Independent Component Analysis), the RR method (Reference Region), and GLCM (Gray-Level Co-occurrence Matrix), to enhance contrast, texture, and detail clarity, further allowing the model to extract more discriminative feature representations. Based on the above inference, it is reasonable to suggest that employing feature enhancement strategies is beneficial for improving performance in BDC. This is also to be expected, as the differences between depth degrees can be subtle, particularly between second- and third-degree burns. Therefore, if image features can be enhanced in clarity, or if models can be guided on what and where to focus when extracting features, they may learn more efficiently, thereby significantly improving accuracy.

**Figure 5.**
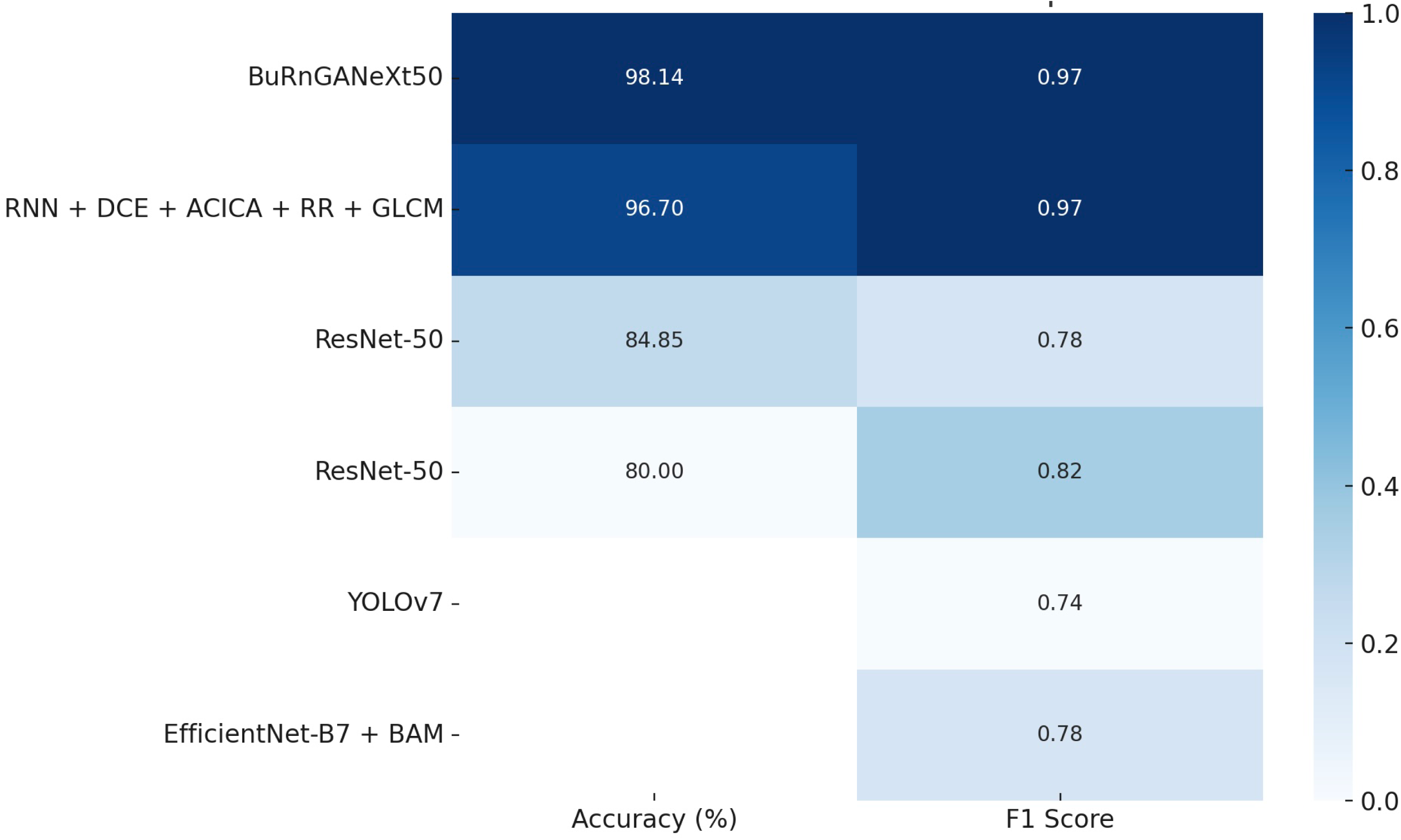
Normalised heatmap of CNN model performance on burn depth classification (BDC), sorted by accuracy. Darker shades indicate higher values of accuracy and F1 score. Models with missing values are shown at the bottom

**Table 3.**
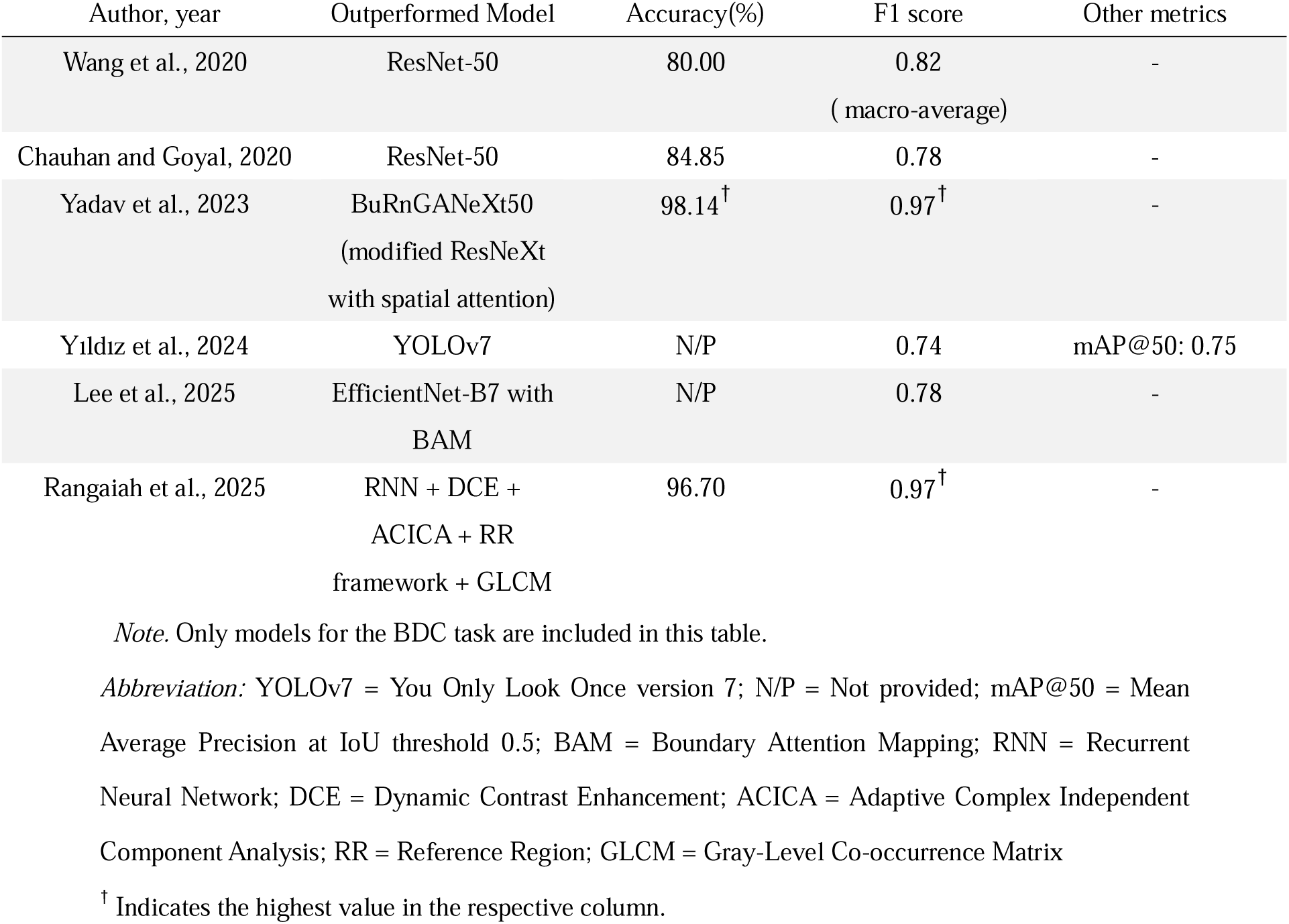
Summary of CNN Models for BDC: Accuracy and Complementary Performance Metrics.

### 4.3 Burn Depth Segmentation

Among the four studies employing BDS, it is challenging to identify a single best-performing architecture, as each study utilised a different CNN model and study design. However, it is worth noting that the model by Cirillo et al. demonstrated notably better performance (Accuracy: 91.89%, DC: 0.92) than that of Liu et al. (Accuracy: 66.84%, DC: 0.68) in BDS, with a 25% higher accuracy and a 0.24 improvement in DC, which was demonstrated clearly in Table 4 and Figure 6. While the two studies adopted different CNN architectures (*Modified U-Net with residual blocks* vs. *HRNetV2-C1*), both models belong to the class of high-performing CNN-based semantic segmentation frameworks widely used in medical imaging. It may be overly simplistic to attribute such a large performance difference solely to the architectural differences. Therefore, it is suggested that the discrepancy may be due to differences in data quality and inconsistent annotations. Cirillo et al. leveraged the TiVi700, a polarised light camera designed to eliminate surface reflections and enhance subdermal contrast, to capture burn images. This approach likely improved the visibility of burn wound features, thereby providing high-quality images for segmentation tasks. In contrast, Liu et al. used standard smartphones and digital cameras, where lighting conditions, reflections, and background noise were likely less controlled in each image, potentially resulting in inconsistent data quality that may have negatively affected the model’s performance. For data annotation, Cirillo et al. defined burn depth degrees based on healing time, which provided a more objective method of annotation. In contrast, Liu et al.’s annotations were based on physicians’ judgement, a method that may involve greater subjectivity and inter-observer variability. It is well-known that high-quality and consistent data annotation is vital for achieving better model performance(38). Taken together, the combination of low-quality input images and inconsistent annotation practices may have led to the notable performance differences observed between the two studies. Therefore, future studies should prioritise the use of high-quality imaging systems and adopt standardised, objective annotation protocols in order to minimise bias and enhance the generalisability of CNN models for BDS.

**Figure 6.**
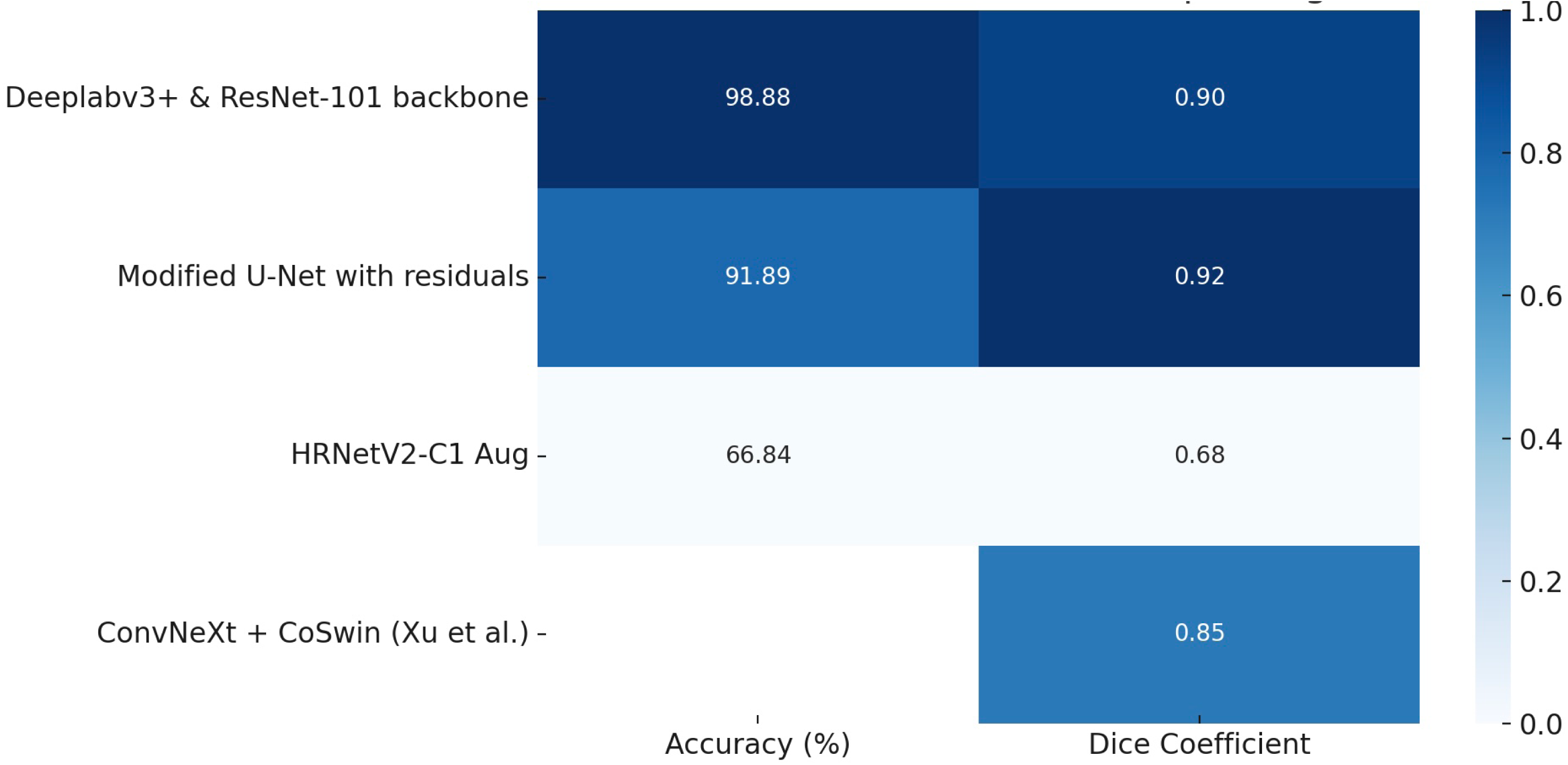
Normalised heatmap of CNN model performance on burn depth segmentation (BDS), sorted by accuracy. Darker shades indicate higher values of accuracy and Dice coefficient. Missing values are shown with gaps.

**Table 4.** Summary of CNN Models for BDS: Accuracy and Complementary Performance Metrics.

| Author, year | Outperformed Model | Accuracy(%) | Dice Coefficient |
| --- | --- | --- | --- |

|  |  |  |  |
| --- | --- | --- | --- |
| Liu et al., 2021 | HRNetV2-C1 Aug | 66.84 | 0.68 |
| Cirillo et al., 2021 | Modified U-Net with residuals | 91.89 | 0.92<sup>†</sup> |
| Chang et al., 2023 | Deeplabv3+ & ResNet-101 backbone | 98.88<sup>†</sup> | 0.90 |
| Xu et al., 2024 | Shared encoder:<br>ConvNeXt + CoSwin<br>BPS Decoder:<br>PointRend( Modified Mask-RCNN)<br>+ FPN<br>BRS Decoder:<br>Convolution layers +DSCU<br>Asymmetric AB | N/P | 0.85 |

*Note.* Only models for the BDS task are included in this table.
*Abbreviation:* BPS = Body Part Segmentation; BRS = Burn Region Segmentation; ConvNeXt = Convolutional Network for Next-generation Vision; CoSwin = Convolutional Shifted Window Self-Attention; DSCU = Dilated Sub-pixel Convolution Upsampling; AB = Attention Block; N/P = Not Provided
<sup>†</sup> Indicates the highest value in the respective column.

As for the remaining two studies included in this prediction task, although the model by Chang et al.(2023) achieved the highest accuracy (98.88%) and the second-highest DC (0.90) in BDS, its study design may partially explain this result. In this study, SLIC (simple linear iterative clustering), a superpixel segmentation preprocessing method, was employed to cluster pixels with similar colour and texture characteristics, thereby facilitating more structured and consistent annotation of deep burn regions by clinicians. This preprocessing step likely reduced annotation noise and contributed to cleaner model inputs. Additionally, the model was only required to distinguish deep burn pixels from areas that had been manually pre-identified as burn regions. In other words, the task was formulated as a binary classification (deep burn vs. other burn wound regions), which is inherently less complex than multi-class prediction. The combination of these factors may have contributed to the model’s high accuracy, although factors such as annotation quality, dataset homogeneity, and model architecture may also have played a role in influencing the results. Xu et al.’s model also achieved a comparably high DC (0.85), despite relying on smartphone- and camera-captured images with physicians-based annotations. This performance may be attributed to its unique architecture, which incorporates a shared encoder for both the Body Part Segmentation and Burn Region Segmentation decoders, an approach that may have positively influenced the model’s performance.

Across all three prediction tasks, model performance is influenced not only by the choice of network architecture but also by the quality of data and annotation. These factors should be carefully considered in future head-to-head comparisons of different CNN architectures.

### 4.4 Potential approaches to improve datasets quality

According to PROBAST+AI, datasets obtained from public image sources or the Google search engine are typically classified as having a high risk of bias due to the lack of systematic and consistent data collection procedures. Additionally, as discussed in Section 4.3, low-resolution images may have significantly and negatively impacted model performance. Nevertheless, acquiring a large volume of high-quality burn images in clinical settings remains challenging, as image collection is time-consuming and requires a diverse and representative patient population. In response to this problem, Dai et al.(39) proposed Burn-GAN, a synthetic image generation framework designed to produce large volumes of annotated burn images for CNN model training. Burn-GAN, derived from StyleGAN, first generates synthetic burn wounds by learning from real burn image distributions, and uses CASC (Color Adjusted Seamless Cloning) to seamlessly embed the generated wound into human skin textures. These synthetic wounds are then mapped onto 3D human models, and automatic annotation is performed in COCO format. Ultimately, Burn-GAN achieved a Frechet Inception Distance (FID) score of 70.97, indicating high image realism, and the CNN model trained on the generated images reached a pixel accuracy of 96.88%. This method may provide a promising approach for CNN-based burn assessment models to improve data quality and reduce both annotation time and inter-observer inconsistency. Nevertheless, important challenges remain in this article. As the study did not report the performance of the CNN model trained exclusively on original images, it is unclear whether the use of GAN-generated data actually improve model performance. Additionally, as diverse real-patient images are still required for realistic synthetic wound generation, the challenge of burn image collection remains an important issue to address. Future research should focus on comparing the performance of models trained on GAN-generated burn images versus real photographs, to validate the generalisability of synthetic datasets in burn wound assessment.

### 4.5 Clinical application of CNN

The potential clinical application of CNN models for burn wound assessment is a central aim of this review. This consideration also underpinned our decision that only studies using RGB photographs were included, as RGB images are readily accessible and offer practical potential for use in both tertiary care hospitals and low-resource settings. Among the fourteen studies, three(23, 27, 28) developed real-time burn wound assessment systems capable of performing automated burn wound segmentation and/or burn depth classification. The models developed by Chang et al. (2023) and Xu et al. further provided TBSA% estimation based on smartphone-captured photographs, along with fluid resuscitation calculations by incorporating patient data such as age and weight. Notably, Chang et al. (2023) and Xu et al. employed different methods for TBSA estimation. While the former used the rule of palms, which required collecting both palm and burn wound images from the patient, the latter utilised the rule of nines, requiring that the burn wound and its corresponding body region (classified by the rule of nines) be included in the image. These applications offer clinicians and specialists a more objective approach to burn wound assessment, facilitating more efficient decisions regarding surgical intervention and improving communication between healthcare providers and patients. However, while these tools demonstrate strong potential, longer-term, evidence-based research is still required to evaluate the consistency between CNN model output and burn specialists’ assessments in burn depth classification and TBSA estimation. Future studies should focus on validating these models in real-world clinical settings over extended periods.

## 5. Conclusion

This systematic review demonstrates that CNN-based models are not only powerful and reliable tools for burn wound assessment, but also capable of delivering outstanding performance when implemented with specific architectures. When applied to burn area segmentation, burn depth classification, and burn depth segmentation, these models have achieved accuracies of up to 98.46%, 98.14%, and 98.88%, respectively, under certain study conditions. This review also identified several meaningful performance patterns across different prediction tasks. For burn area segmentation, the combination of *ResNet-101 and ASPP* appears to provide a strong and reliable baseline, while *HRNetV2-C1* can also play an important role when low-resolution data are used or when precise spatial detail is required. For burn depth classification, feature enhancement strategies—such as spatial and channel attention modules, as well as image preprocessing methods—were observed to likely improve model performance. In the case of burn depth segmentation, high-quality data and consistent image annotation appear to play a vital role and may substantially improve model performance. Taken together, both network architectures and the quality of data, including image resolution and annotation consistency, collectively influence CNN model performance.

This review has certain limitations. Due to the heterogeneity across studies, the interpretation of findings should be approached with caution. Direct head-to-head comparisons across different CNN models should be conducted in future research to more accurately identify optimal architectures and hyperparameters for each prediction task.

Despite these challenges, this review provides valuable insights into the selection of CNN architectures for burn wound assessment. It also underscores the reliability and potential of CNN models trained on RGB photographs, supporting the future integration of such models into mobile phone–based applications. Future research can build upon these findings to develop real-time mobile tools capable of assessing burn area and burn depth, alongside total body surface area (TBSA) estimation, thereby enabling accurate and immediate comprehensive assessment for patients.

## Data Availability

All data produced in the present work are contained in the manuscript

## 5.3 Ethical approval

No ethical approval was required for this study.

## 5.4 Data Availability Statement

No new data were generated or analysed in this study. Data sharing is not applicable to this article.

