## Supplementary Table 1 for "Convolutional Neural Networks for Burn Segmentation and Classification Tasks Using RGB Photographs: A Five-Year Systematic Review"

### Supplementary Table 1 : Risk of Bias using PROBAST+AI Tool

| Study | Type | D1 | D2 | D3 | D4 | Overall RoB | Applicability |
|---|---|---|---|---|---|---|---|
| Wang et al. (2020) | Development | High | Low | Unclear | High | High | Some concerns |
| Chauhan & Goyal (2020) | Dev + Val | High | Low | High | Unclear | High | High concerns |
| Liu et al. (2021) | Dev + Val | Low | Low | Unclear | Low | Moderate | Some concerns |
| Chang et al. (2021) | Dev + Val | Low | Low | Low | Low | Low | Some concerns |
| Chauhan & Goyal (2021) | Dev + Val | High | Low | Unclear | Low | High | Some concerns |
| Cirillo et al. (2021) | Dev + Val | Low | Low | Low | Low | Low | Some concerns |
| Chang et al. (2023) | Dev + Val | Low | Low | Low | Low | Low | Low |
| Boissin et al. (2023) | Dev + Val | Low | Unclear | Unclear | Low | Moderate | Some concerns |
| Yadav et al. (2023) | Dev + Val | High | Low | Unclear | Low | High | Some concerns |
| Jacobson et al. (2023) | Dev + Val | High | Low | Low | Low | High | Some concerns |
| Yıldız et al. (2024) | Dev + Val | High | Low | Unclear | Low | High | Some concerns |
| Xu et al. (2024) | Dev + Val | Unclear | Low | Low | Low | Moderate | Some concerns |
| Lee et al. (2024) | Dev + Val | Low | Low | Unclear | Low | Moderate | Some concerns |
| Rangaiah et al. (2025) | Dev + Val | Low | Low | Low | Low | Low | Low |
