## Supplementary Table 3 for "Convolutional Neural Networks for Burn Segmentation and Classification Tasks Using RGB Photographs: A Five-Year Systematic Review"

### Supplementary Table 3: Studies excluded after full-text review with reasons

| Study (Author, Year) | Reason for Exclusion |
|---|---|
| Chen et al., 2024 | Study focused on CNN adversarial training rather than segmentation or classification tasks |
| Rambhatla et al., 2021 | Outcome not related to burn segmentation or classification. |
